# Antidiabetic Activity of a Ghanaian Herbal Product, DBT-57A: An Observational Study

**DOI:** 10.1101/2025.05.14.25327662

**Authors:** Rudolph Mensah, Kwesi Prah Thomford, Abraham Yeboah Mensah, Isaac K. Amponsah, Ama Kyeraa Thomford, Gustav Komlaga

## Abstract

**Introduction:** The rising global burden of diabetes mellitus has led to growing interest in herbal therapies as alternative or complementary treatments. In many low-resource settings, herbal medicines remain a cornerstone of diabetes management. DBT-57A, a Ghanaian polyherbal formulation, has been used in routine community health practice for over three decades without formal scientific evaluation. This study assessed its real-world clinical effectiveness and safety among patients with type 2 diabetes mellitus (T2DM).

**Methods:** A 28-day prospective non-randomized observational study was conducted involving 25 adults with clinically diagnosed T2DM who were already receiving DBT-57A as part of routine care at a Ghanaian health facility. Fasting blood sugar (FBS) levels were recorded at baseline, Day 14, and Day 28. Diabetes-related symptoms (polyuria, nocturia, weight loss) and any adverse events were documented through patient interviews. Statistical analysis was performed using a paired t-test to compare FBS levels at baseline and at Days 14 and 28. Ethical approval was obtained from the Committee on Human Research, Publications and Ethics at KNUST/Komfo Anokye Teaching Hospital.

**Results:** Mean FBS levels decreased significantly by 49.8%, from 18.34 ± 6.54 mmol/L at baseline to 9.19 ± 3.76 mmol/L on Day 28 (p < 0.05). Symptomatic relief was reported by 95.2% of participants. No serious adverse events occurred.

**Conclusion:** DBT-57A showed promising glucose-lowering effects and symptom improvement in this observational cohort. These findings support further investigation through randomized controlled trials to confirm its efficacy, ensure safety, and explore its role in integrative diabetes care in low-resource settings.

## 1.0 Introduction

### 1.1 Background

Diabetes mellitus (DM) is a prevalent metabolic disorder characterized by chronic hyperglycemia resulting from relative insulin deficiency, resistance, or a combination of both factors (1). Diabetes can be broadly classified into two types, type I (insulin-dependent) and type II (non-insulin-dependent) (2). Globally, the incidence of diabetes is rapidly increasing, paralleling the rise of other lifestyle-related diseases like hypertension (3,4). In 2021, approximately 529 million people were living with type 2 diabetes, projected to exceed 1.31 billion by 2050 (5). The economic burden was estimated at $1.3 trillion in 2015, with an expected rise to $2.1 trillion by 2030 (6). These alarming trends make diabetes a critical global public health concern, demanding effective management strategies including dietary changes, pharmacological treatment and lifestyle modifications (7).

In Ghana, despite the availability of conventional medical treatments for diabetes and its associated complications, many patients turn to herbal medicine for chronic disease management. This preference is often due to the perception that herbal medicines are not only safer but efficacious (8). In many rural communities (9–11), herbal medicine remains a primary therapeutic option due to limited access to conventional care (12–16).

Numerous medicinal plants with antidiabetic potential have been documented in the literature (13–15,17). Many plant-based products have been identified for their potential antidiabetic properties, leading to the incorporation of medicinal plants into various treatment models, either individually or in combination (18,19). In Ghana, such plant-based medicinal products, including polyherbal supplements, are tested for quality and safety by accredited research and testing laboratories in various institutions. A number of these products are approved by the Food and Drugs Authority (FDA) and widely marketed as nutritional supplements for blood sugar modulation, despite the lack of valid clinical data.

Conversely, several herbal products are dispensed solely within prescribing facilities, exempting them from regulatory oversight due to their extemporaneous nature. This regulatory gap introduces risks to patients and undermines the credibility of herbal medicines in primary healthcare, especially when products fail to meet standardized quality, safety, and efficacy criteria. DBT-57A is one such polyherbal product that has been used for over three decades in an underserved Ghanaian community where many diabetic patients rely on it for glycemic control. Although clinicians have reported patient improvement, the product lacks clinical validation.

DBT-57A is composed of *Solanum torvum, Allium cepa*, and *Khaya senegalensis,* plants traditionally used for diabetes management. Preclinical studies have demonstrated their hypoglycemic, antioxidant, and anti-inflammatory properties. Detailed active constituent profiles are provided in Supplementary Information (S1 Table).

Despite their prospects, the growing reliance on herbal remedies for chronic disease management despite limited clinical validation highlights an urgent need for evidence-based integration of traditional medicines into primary healthcare systems.

Given the economic burden of diabetes, patient preferences for alternative treatments, and the long-standing, informal clinical use of DBT-57A, this study aimed to conduct a prospective observational evaluation of its real-world effectiveness and safety in patients with type 2 diabetes (9,20). Conducted in a community health facility where the product was already in routine use, the study assessed fasting blood sugar reduction, symptom improvement, and tolerability under naturalistic conditions.

Specifically, we assessed changes in fasting blood sugar (FBS) by Day 28, symptom relief by Days 14 and 28, safety through adverse event monitoring, and the relationship between dosage and clinical response. These objectives were largely met, with findings supporting future formal evaluation through randomized trials. This study aims to assess whether DBT-57A can significantly reduce fasting blood sugar (FBS) levels and alleviate diabetes-related symptoms in patients with type 2 diabetes mellitus.

## 2.0 Materials and Methods

### 2.1 Test Product (DBT-57A)

DBT-57A is a polyherbal decoction produced with *Solanum torvum*, *Allium cepa*, and *Khaya senegalensis* with established use for the management of diabetes for over three decades. Patients attending the Health for All Herbal Clinic in Asamankese, Eastern Region of Ghana, are prescribed DBT-57A as part of their routine diabetes care. The prescribed dosage is 80 mL, taken three times daily, for a total monthly supply of 6 L. Basic quality and safety tests of DBT-57A were conducted under a reverse pharmacology framework to ensure its suitability for observational evaluation. Results of phytochemical, microbial, physicochemical, and heavy metal analyses are provided in Supplementary File S1 Table.

### 2.2 Study Design

This was a non-randomized, non-interventional observational study. The primary aim was to assess the association between use of DBT-57A and changes in fasting blood sugar and diabetes-related symptoms. The study was not designed to establish causal relationships, and no control group or blinding was employed.

#### 2.2.1 Target Population and Sample Size

Patients visiting the Health facility with clinically established diabetes symptoms such as polydipsia, polyuria, nocturia, blurred vision, lethargy and weight loss were recruited for observation. Again, known diabetics who had defaulted on their conventional treatment for over a month were recruited for inclusion after satisfying the study criteria. A convenience sample of 25 participants was determined based on the average number of diabetes cases seen annually at the facility, consistent with feasibility-based approaches typical in observational studies.

#### 2.2.2 Inclusion Criteria and Exclusion Criteria

Inclusion: Males and females aged 20–85 years with a diagnosis of T2DM and FBS > 7.0 mol/L after 8 hours of fasting.

Exclusion: Type 1 diabetes mellitus, insulin use, cardiovascular/renal/hepatic diseases, pregnant/breastfeeding women, and individuals using treatments contraindicated with herbal medicines.

#### 2.2.3 Recruitment of Participants

This study was conducted over a six-month period, from September 2019 to February 2020, as an outpatient investigation. As part of the usual clinical intake, patients were offered the opportunity to participate in the observational monitoring of their response to DBT-57A. No new intervention was introduced. Participants were recruited prospectively, with data collected on every other new case of diabetes mellitus. Each participant was monitored for 28 days, during which fasting blood sugar levels were measured. Sampling took place Monday to Saturday between 7:30 AM and 10:00 AM. Newly diagnosed diabetic patients were recruited for the study and instructed to repeat their FBS after an 8-hour overnight fast. The purpose of the study, along with patient anonymity and rights, were explained to them in English or a local Ghanaian language. Informed consent was obtained from all participants prior to study participation, as detailed in the Ethical Considerations section. Upon consenting, participants completed a structured questionnaire that gathered data on their demographics (contact details, age, occupation, gender, educational level), body weight, blood pressure, and clinical symptoms related to diabetes. A similar recruitment protocol was employed for known diabetics who had defaulted on their conventional therapy for a period of one month.

### 2.3 Classification of Efficacy

Clinical assessments, including patient history, physical exams, and lab tests, were conducted on days 0, 14, and 28. Interim phone follow-ups were conducted on days 1, 3, 18, and 21 monitored compliance, treatment progress, and adverse reactions, while in-person follow-ups on days 14 and 28 included assessment of vital signs, weight, dietary compliance and newly reported complaints.

#### 2.3.1 Primary Outcome Measure

The primary outcome measure for evaluating the clinical efficacy of DBT-57A was the reduction in fasting blood sugar (FBS) levels by day 28.

Based on the FBS values at the end of the study, participants were classified into three categories. An effective response was defined as achieving FBS levels (FBS 4.4-6.7 mmol/L by Day 28), partial response (FBS 6.7-10.7 mmol/L at the end of the study period. A response was considered ineffective when there was less than a 30% reduction in FBS levels from baseline, no observable clinical improvement or worsening condition.

#### 2.3.2 Secondary Outcome Measure

Secondary outcome measures focused on clinical improvements in diabetes-related symptoms and changes in blood sugar levels. Specifically, improvements in symptoms such as polyuria, nocturia, and weight loss were assessed on day 14 and at the end of the study. In addition, the percentage reduction in fasting blood sugar from baseline to day 28 was calculated for each participant to further support the evaluation of treatment response.

### 2.4 Ethical Considerations

This study was a non-interventional, observational investigation conducted at a local herbal clinic where DBT-57A has been used as part of routine diabetes management for over 30 years. Participants were not randomized or prospectively assigned to DBT-57A by the investigators; instead, they were individuals already receiving the product as part of their usual care. The aim of this study was to observe and document outcomes associated with this long-established treatment to inform future research.

The study received ethical approval from the Committee on Human Research, Publications and Ethics (CHRPE) at the School of Medical Sciences, Kwame Nkrumah University of Science and Technology/Komfo Anokye Teaching Hospital (Reference Number: SMS/KATH CHRPE/AP/0548/19) in September 2019. Institutional approval was also obtained from the local health facility where the study was conducted. The study adhered to Good Clinical Practice guidelines.

Informed written consent was obtained from all participants prior to any study-related procedures. Participants received a detailed information sheet explaining the study’s purpose, procedures, potential risks, and their rights. The consent form and participant information sheet were provided in both English and a local Ghanaian language where needed. Participants had adequate time to review the documents and ask questions before signing. A research staff member witnessed the process to ensure validity. No minors were included, and consent was not waived by the ethics committee. The questionnaire, participant information sheet, and consent form are provided as supplementary materials (S2 Text, S3 Text, and S4 Text respectively).

### 2.5 Data Collection and Statistical Analysis

Data collected was entered into Microsoft Excel. Data analysis was conducted using Stata/MP 17. Descriptive statistics were used to summarize participant demographics and baseline characteristics, and paired t-tests were employed to compare FBS values from baseline to Day 28. Results were expressed as mean ± SD [95% CI] and statistical significance was set at *p* ≤0.05.

## 3.0 Results

### 3.1. Demographics of Participants

The study included 25 participants, with 15 (60.0 %) females and 10 (40.0 %) males. Four participants were lost to follow-up, resulting in 21 completing the 28-day study, yielding a participant completion rate of 84.0% (Table 3.1). The majority (76.20%) were aged between 40 and 60 years representing the most productive age bracket. Participants aged 80 and above were the least represented (4.76%) (Table 3.2).

**Table 3.1.** Summary of demographic data for participants in the study.

| **Gender** | ***Recruited Participants*** | ***Completed the study*** |
| --- | --- | --- |
| Male | 10 (40.0) | 9 (90.0) |
| Female | 15 (60.0) | 12 (80.0) |
| Total | 25 (100.0) | 21 (84.0) |

Data presented as n (%)

**Table 3.2.**
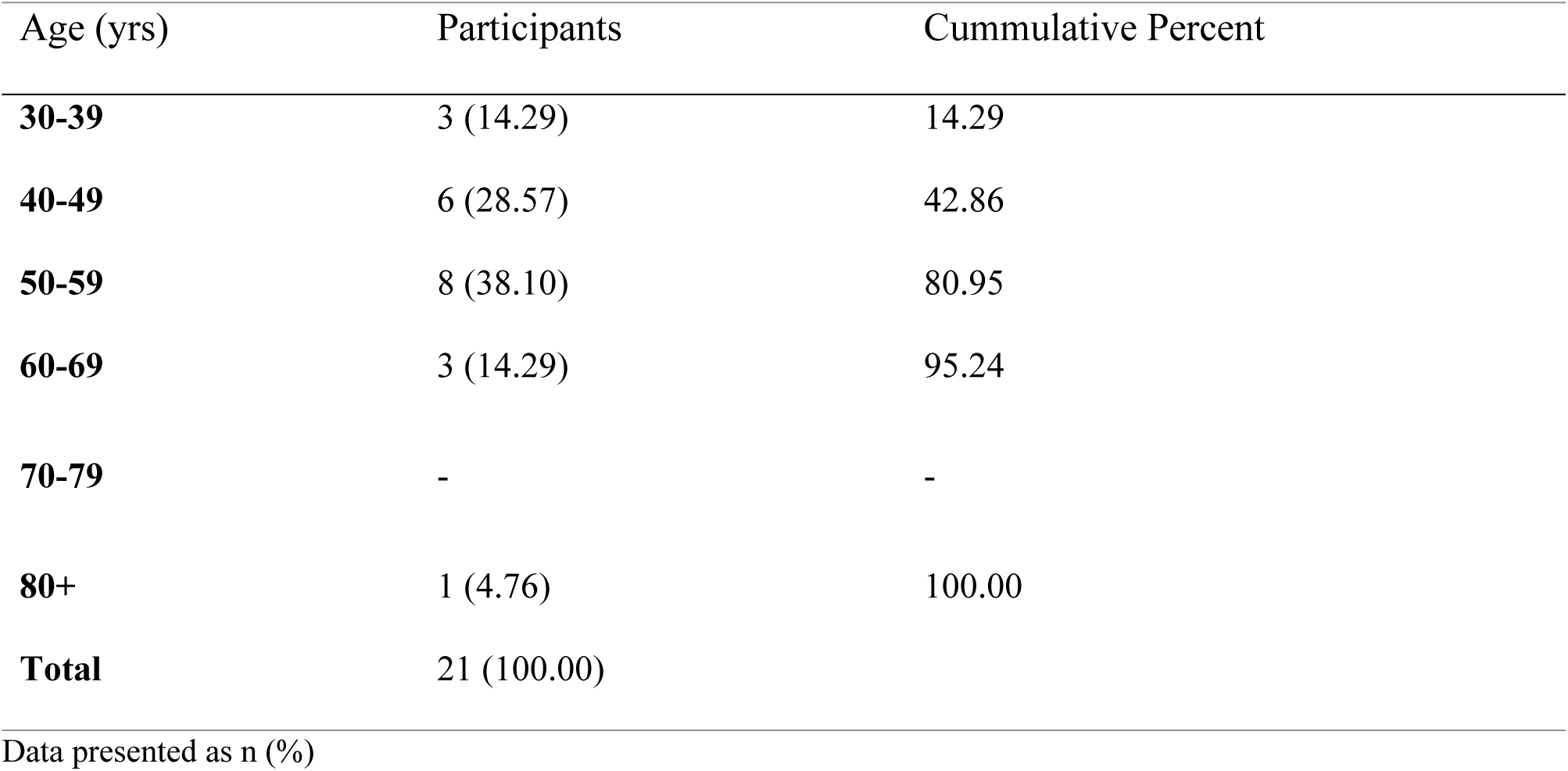
Summary of age group demography for participants in the study.

### 3.2 Treatment Efficacy of DBT-57A

#### 3.2.1 Effects of DBT-57A on blood sugar levels

Overall, the blood sugar level of participants decreased significantly from 18.33 mmol/L before treatment to 9.2 mmol/L at the end of the study, with a mean change of -9.13 mmol/L. During the treatment, a 6.05 mmol/L reduction in fasting blood sugar (FBS) was observed on day 14, which further decreased by 3.10 mmol/L by day 28. Statistically significant changes were noted between day 0 and both day 14 and day 28 (Table 3.3 & Fig 1). 8 patients who had high initial FBS levels (20 mmol/L up to 33.3 mmol/L) improved over time. Out of 21 participants, 10 (47.62%) achieved the primary outcome of effective clinical response, 6 (28.57%) had partial response, and 5 (23.81%) had no response (Table 3.4). A statistically significant reduction in FBS was observed on both Day 14 and Day 28 (p < 0.05), supporting the treatment’s efficacy.

**Table 3.3.** Mean FBS and paired *t*-test analysis of FBS on Days 0, 14 and 28.

| FBS | Mean FBS [95% CI] | T-test | Mean Diff ± SD | p-value ($\alpha = 0.05$) |
| --- | --- | --- | --- | --- |
| **Day 0** | 18.34 ± 6.54<br>[15.36 – 21.32] | fbs_day0 vs<br>fbs_day14 | 6.05 ± 1.02 | <0.0001 |
| **Day 14** | 12.29 ± 5.39<br>[9.83 – 14.74] | fbs_day14<br>vs<br>fbs_day28 | 3.1 ± 4.01 | <0.0021 |
| **Day 28** | 9.19 ± 3.76<br>[7.47 – 10.90] | fbs_day 0 vs<br>fbs_day28 | 9.15 ± 5.57 | <0.0001 |

Values represent Mean ± SD [95% CI] for participants (n = 21); p <0.05 (paired *t*-test comparing the means to baseline).

**Table 3.4.** Participants attaining the study criteria of clinical response Clinical response n (%)

| Clinical response | n (%) |
| --- | --- |
| Efficacy | 10 (47.62) |
| Partial efficacy | 6 (28.57) |
| No efficacy | 5 (23.81) |
| Total | 21 |

**Fig 1.**
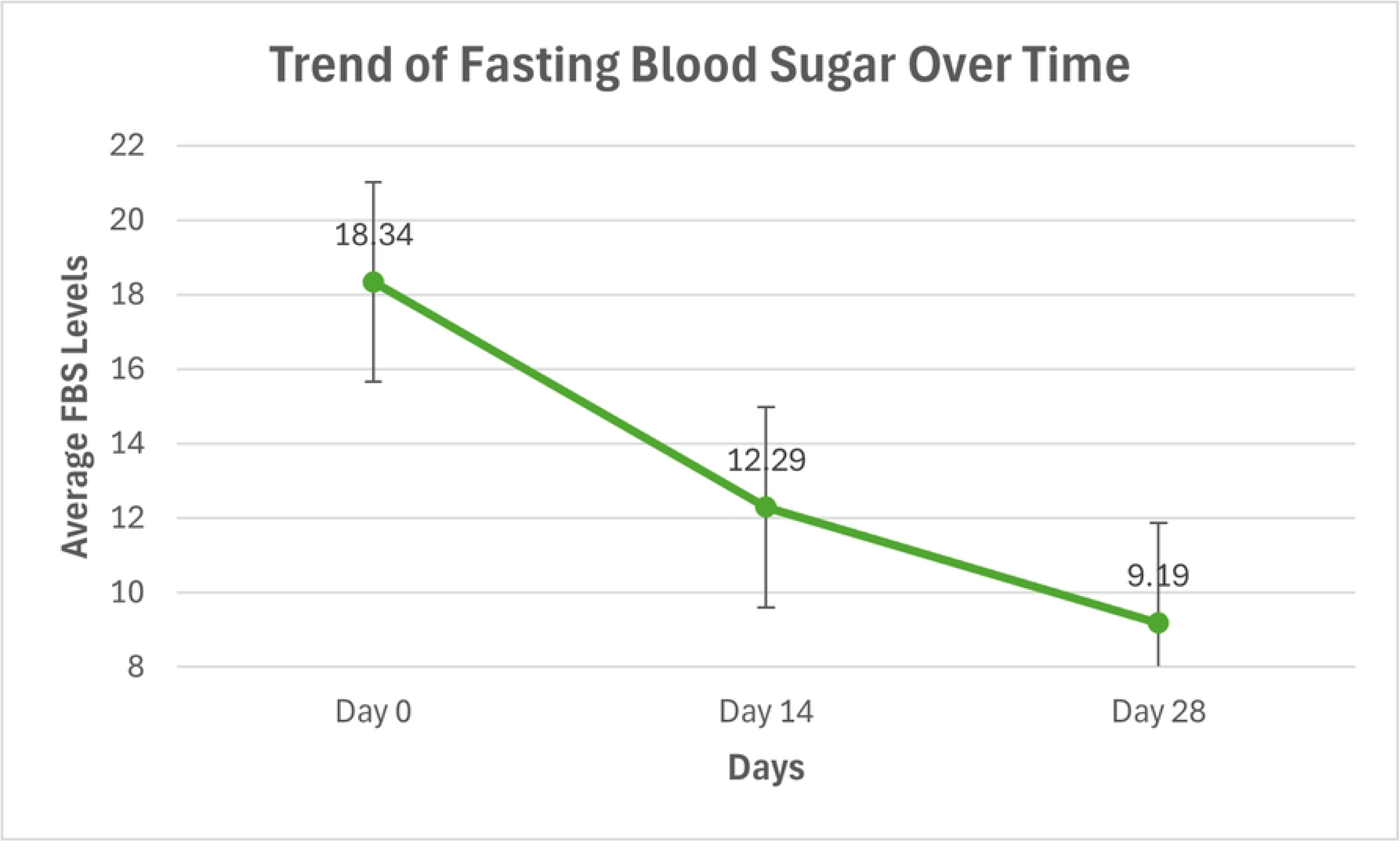
Trend of Fasting Blood Sugar Over Time. *A line graph showing the change in mean fasting blood sugar (FBS) levels among participants from baseline through Day 28 of DBT-57A administration*.

The Boxplot shows a clear decrease in median FBS levels over time (Fig 3.2). The spread (interquartile range) reduces from Day 0 to Day 28, showing more consistency in values by the end. A few outliers appear, indicating some patients had significantly higher or lower FBS than the group.

**Fig 2.**
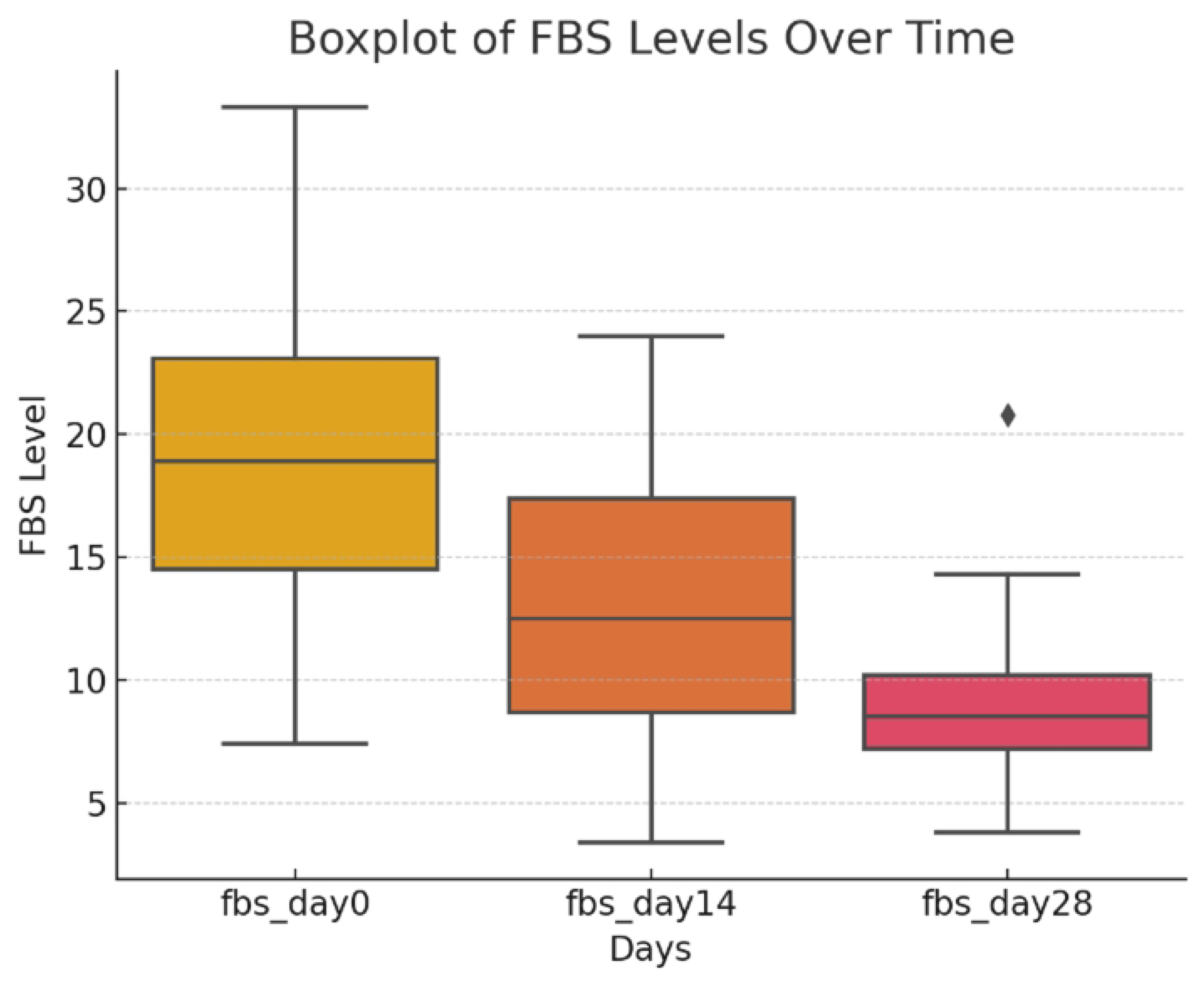
Fasting Blood Sugar Distribution During the Study. *A box plot illustrating changes in fasting blood sugar (FBS) levels among participants from baseline through follow-up after DBT-57A administration. Significant reductions in FBS were observed (P < 0.01; *P < 0.001) compared to baseline values.*

##### 3.2.2 FBS Reduction by Age Group

All age groups showed improvement over time. The 40-49 and 50-59 age groups started with high FBS levels but saw significant drops. The 60-69 age group had the lowest FBS by Day 28.

**Table 3.5.**
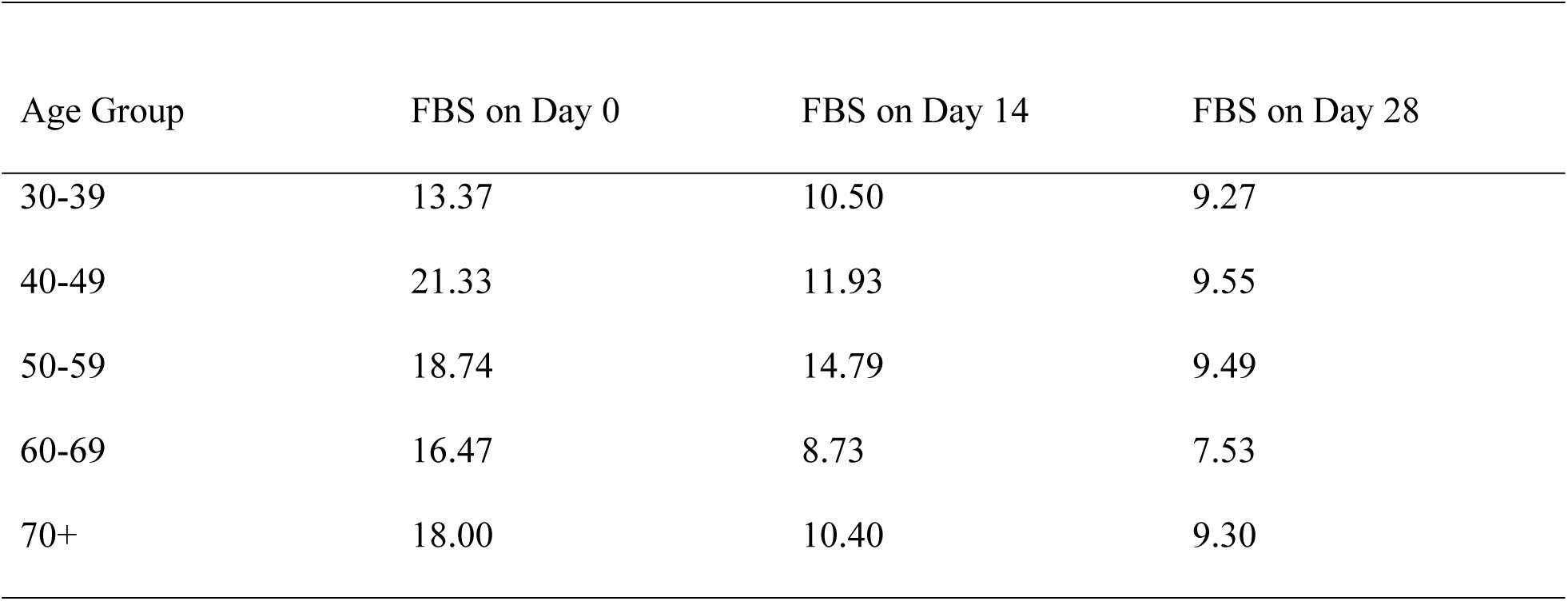
FBS Reduction by Age Group.

The correlation analysis between age and fasting blood sugar (FBS) levels at different time points (Day 0, Day 14, and Day 28) showed weak correlations (Table 3.6). The Pearson correlation coefficient for Age and FBS on Day 0 was r = 0.10, for Day 14, r = -0.01, and for Day 28, r = - 0.01. These values suggest that there is no strong relationship between age and fasting blood sugar levels at any of the time points. However, there were strong positive correlations (0.71 between Day 0 and Day 14), indicating that patients with high initial FBS tend to have higher levels throughout the study.

**Table 3.6.** Correlation between Age and FBS levels on Day 0, 14 and 28.

|  | Age | fbs_day0 | fbs_d~14 | fbs_d~28 |
| --- | --- | --- | --- | --- |
| Age | 1.0000 |  |  |  |
| fbs_day0 | 0.1013<br>0.6622 | 1.0000 |  |  |
| fbs_day14 | -0.0129<br>0.9556 | 0.7112<br>0.0003 | 1.0000 |  |
| fbs_day28 | -0.0112<br>0.9617 | 0.5269<br>0.0141 | 0.6680<br>0.0009 | 1.0000 |

##### 3.2.2 Participants Blood Sugar Level Per Visit

A line graph with markers illustrating individual participants’ fasting blood sugar (FBS) levels on each day of visit (Figure 3.3) demonstrates the variation in blood glucose trends across individuals. Additionally, the mean FBS for all participants on each visit day (Figure 3.2) exhibits a significant decline over time, indicating an overall improvement in blood glucose control throughout the study period.

**Fig 3.**
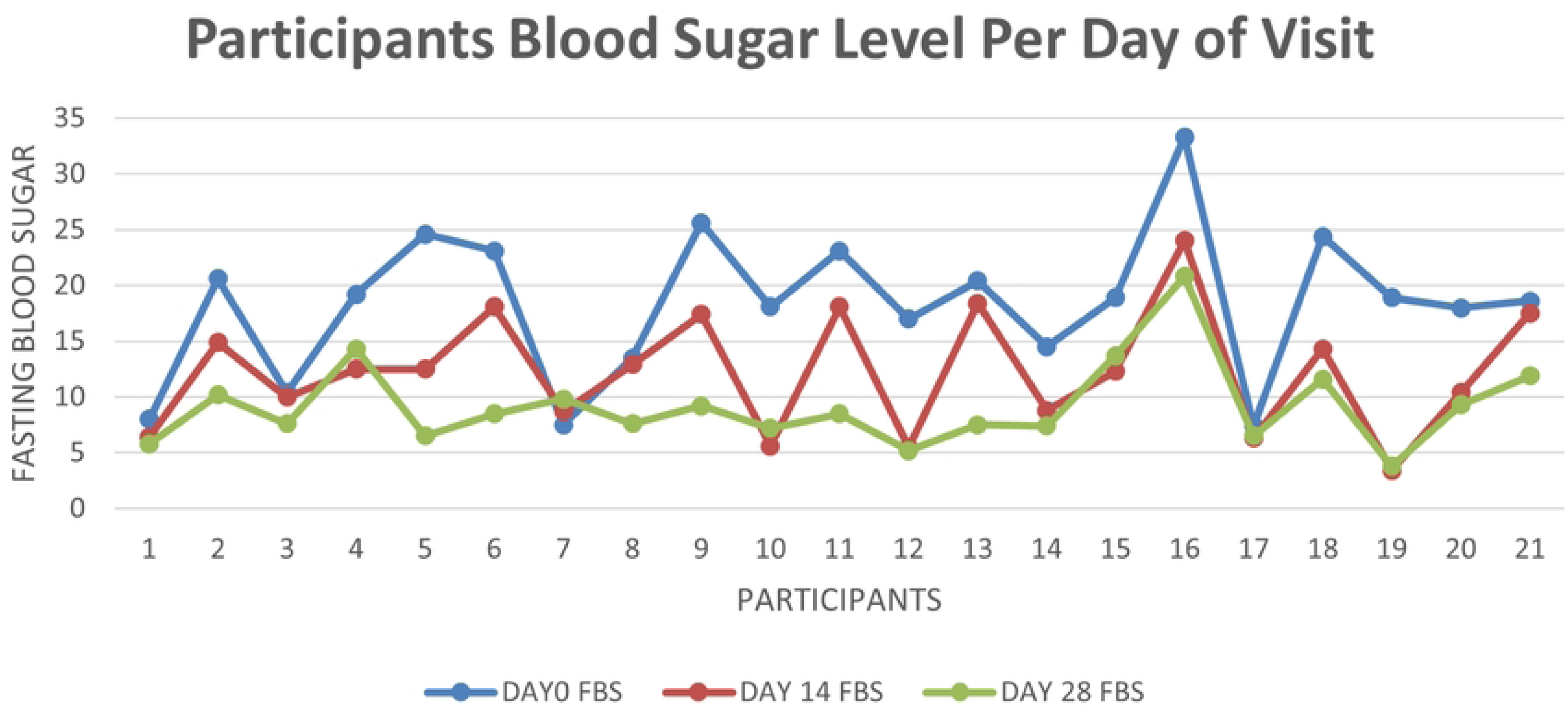
Participants’ Blood Sugar Levels by Day of Visit. *A grouped chart displaying individual fasting blood sugar (FBS) values recorded per participant at each scheduled visit. The figure highlights variation in glycemic response to DBT-57A during the 28-day study period*.

**Fig 4.**
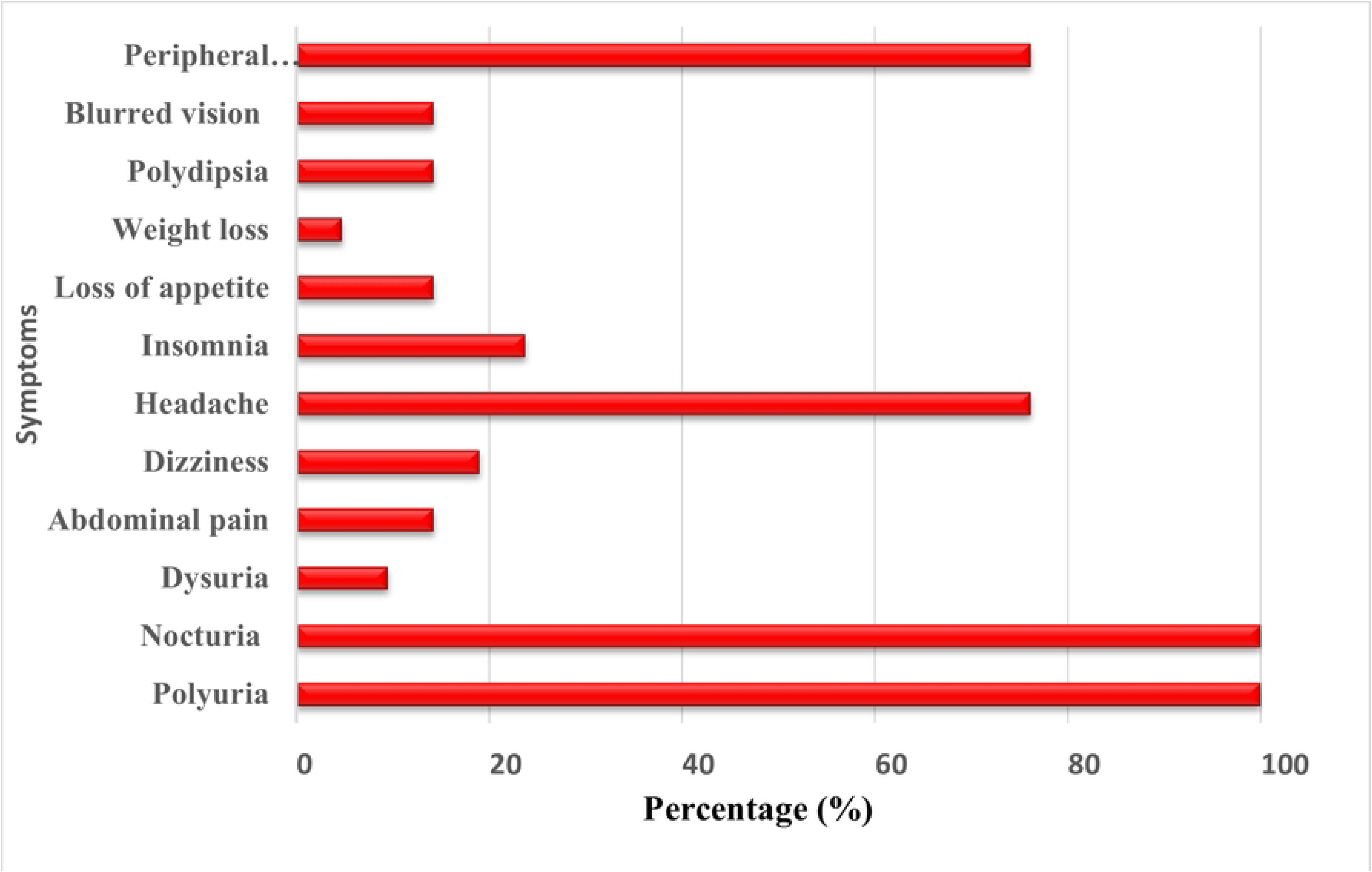
Diabetes-Related Symptoms Reported by Participants. *A bar chart summarizes the frequency of self-reported diabetes-related symptoms such as polyuria, nocturia, and weight loss before and after DBT-57A administration. The figure illustrates changes in symptom prevalence during the study period*.

#### 3.3. Secondary Outcome

##### 3.3.1 Body Weight

The change in participants’ mean body weight from 64.67 kg at baseline to 64.48 kg at the end of the study was not statistically significant (Table 3.5), indicating minimal variation in weight over the study period.

**Table 3.7.** Body Weight Assessment of Participants Before and After Treatment.

| Parameter | Before Treatment | After Treatment |
| --- | --- | --- |
|  | (Mean ± SD) | (Mean ± SD) |
| Body Weight (kg) | 64.67 ±14.43 | 64.48 ± 13.56 |

Values represent Mean ± SD for diabetic patients (n=21).

##### 3.3.2 Blood Pressure

Blood pressure levels exhibited a slight decline as the treatment progressed; however, the change was not statistically significant (Table 3.8), suggesting that the treatment had a minimal effect on blood pressure over the study period. Despite a slight reduction in body weight and blood pressure, these changes were not statistically significant (p > 0.05), suggesting that DBT-57A’s primary benefit lies in glucose control rather than weight or blood pressure management.”

**Table 3.8.** Blood Pressure Assessment.

| Parameter | Day 0 | Day 14 | Day 28 |
| --- | --- | --- | --- |
|  | Mean | Mean | Mean |
| Blood Pressure (Systolic mmHg) | 123 ± 26.30<sup>1</sup> | 121 ± 19.47 | 122 ± 18.61 |
| Blood Pressure (Diastolic mmHg) | 79 ± 15.58 | 75 ± 12.09 | 77 ± 10.17 |

<sup>1</sup>Values represent Mean ± SD for participants (n=21).

##### 3.3.3. Improvement of Symptoms of Diabetes Mellitus presented by Participants

On Day 0, all participants presented with polyuria and nocturia, while peripheral numbness was the third most reported symptom. Additional symptoms included headache, insomnia, blurred vision, weight loss, dysuria, dizziness, and polydipsia.

##### 3.3.4 Symptoms Improvement from Day 14 to Day 28

Between Day 14 and Day 28, all symptoms showed further improvement, with many achieving complete resolution. By Day 28, polyuria, nocturia, dysuria, dizziness, insomnia, weight loss, polydipsia, blurred vision, peripheral numbness, and burning sensation had 100% improvement, while headache, abdominal pain, and general body pains showed continued but slightly slower progress. Loss of appetite, which had no improvement at Day 14, reached full recovery by Day 28, indicating overall positive symptom resolution.

**Table 3.9.**
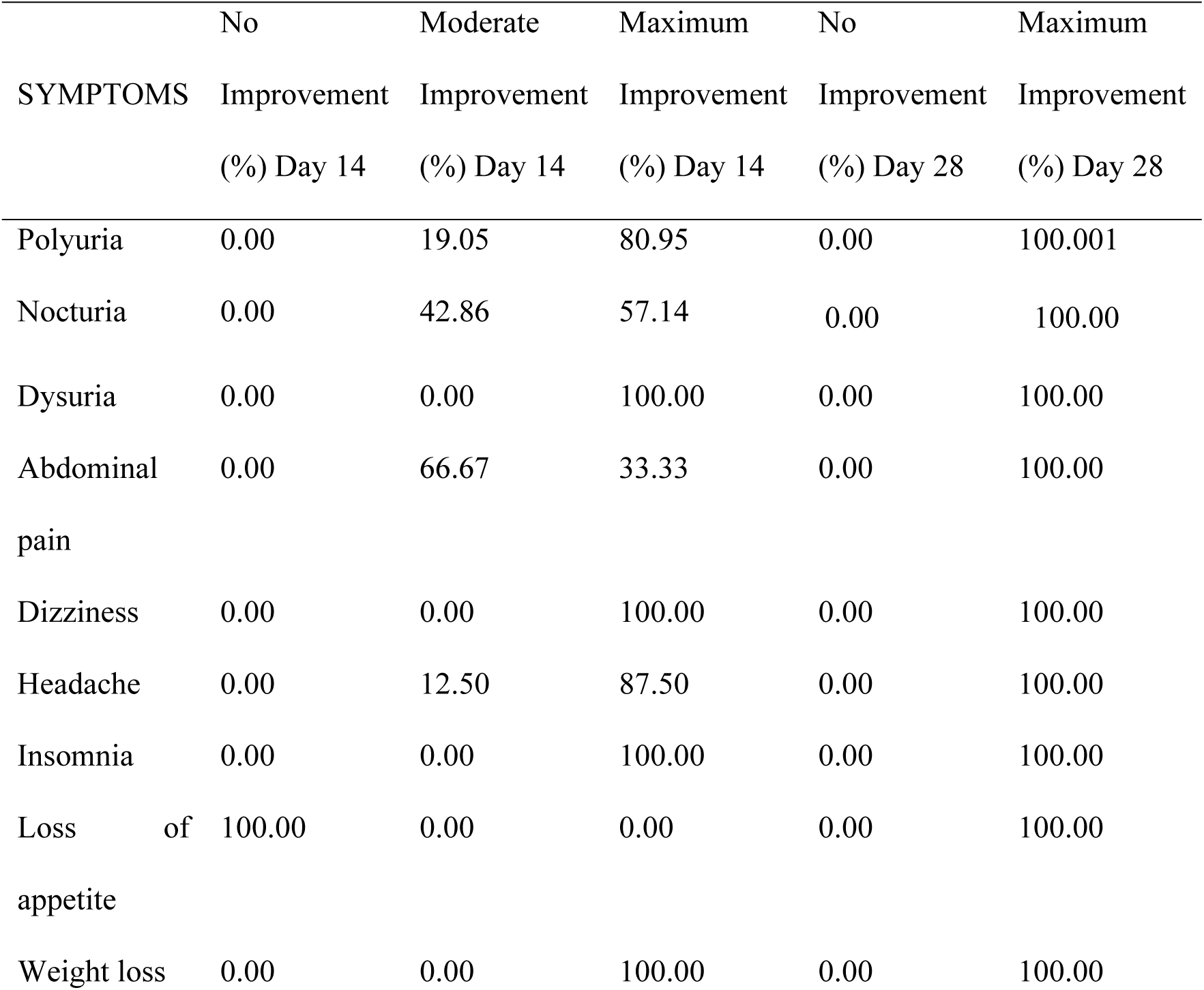
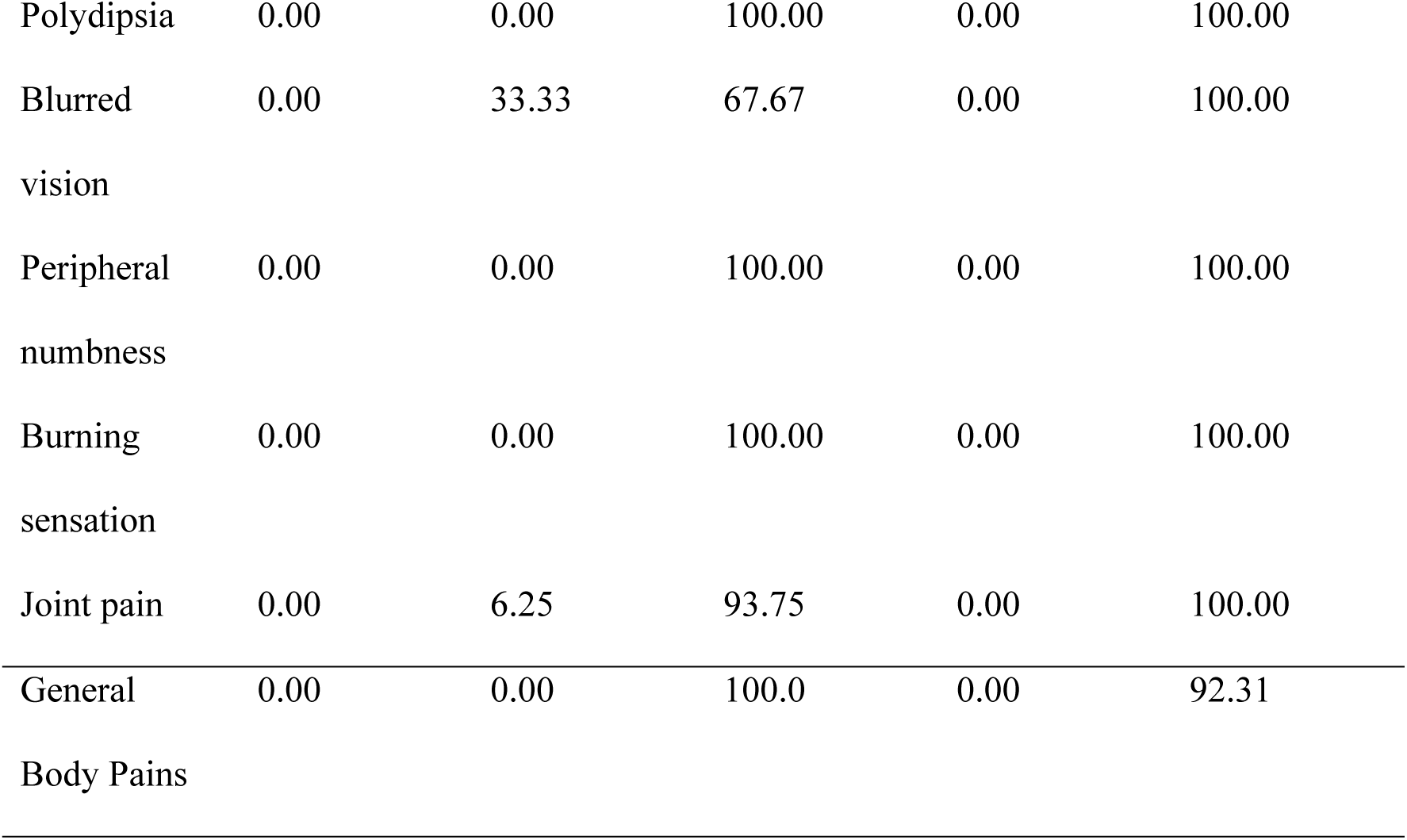
Symptom Improvement from Day 14 to Day 28.

##### 3.3.5. Medical Decision Based on Clinical Outlook of Participants using DBT-57A

All participants exhibited symptom improvement, particularly within the first week of treatment. By the end of the study, polyuria, nocturia, and weight loss showed 100% resolution, and there was an overall 95.24% improvement in diabetic condition, as evidenced by a decline in fasting blood sugar (FBS) levels. However, one participant (4.76%) experienced an increase in FBS from baseline by Day 14 and showed no improvement by the end of the study. Based on clinical assessment, all 20 improved participants were not referred for further treatment, while the one non-improving participant also did not receive a referral (Table 3.10).

**Table 3.10.** Clinical Condition and Medical Decision of Participants who took *DBT -57A*.

| Clinical Condition | Medical Decision |  |  |
| --- | --- | --- | --- |
|  | Not referred | Referred | Total |
| Improved | 20 | 0 | 20 |
| No Change | 1 | 0 | 1 |
| **Total** | 21 | 0 | 21 |

#### 3.4 Safety and Tolerability

No serious adverse events were reported during the study period. The overall tolerability of DBT-57A was high, with no participants discontinuing treatment due to adverse reactions. Mild and self-limiting adverse effects such as diarrhea and dizziness were observed in 2 of the participants, but these did not require medical intervention.

### 4.0 Discussion

#### 4.1 Study Findings

Herbal medicines have gained widespread acceptance over the past three decades, due to their accessibility and traditional use in treating various ailments. However, ensuring their safety and efficacy remains a challenge (21). Potential risks such as hepatotoxicity, nephrotoxicity, cardiovascular complications, and herb-drug interactions, especially when combined with antidiabetic medications, are of concern. (22,23).

The lack of regulation, variability in preparation methods, and unmonitored use alongside conventional treatments highlight the need for public education and research on drug-herb interactions. Herbal formulations like DBT-57A, which show potential as adjunct therapies for diabetes management, require rigorous safety analysis and standardized testing to establish clear safety parameters (9,24,25).

Our findings suggest that DBT-57A may be a promising adjunct therapy for diabetes management, demonstrating a mean fasting blood sugar (FBS) reduction of 9.13 mmol/L over 28 days, with most improvement occurring within the first two weeks. This observed hypoglycemic activity suggests a potential early pharmacodynamic effect. However, further research is needed to assess whether this reduction stabilizes or continues beyond 28 days, and to elucidate the underlying mechanisms of action.

Although early pharmacodynamic effects were observed, no significant correlation was found between age and fasting blood sugar (FBS) levels. This finding aligns with previous studies suggesting that glycemic control is influenced more by factors such as lifestyle, diet, and medical history rather than age alone (26,27)

However, variability in response (with 23.81% of participants showing no response) suggests that other factors, such as genetics, disease severity, or adherence to treatment, may play a role in treatment efficacy. Future studies should explore biomarkers or patient characteristics that predict treatment responsiveness.

Despite the significant glycemic improvements, no meaningful changes in body weight or blood pressure were observed during the study. This suggests that DBT-57A primarily targets blood sugar regulation, without significantly affecting other metabolic parameters. However, diabetes-related symptoms such as polyuria, nocturia, and peripheral numbness showed high resolution rates of resolution, with some achieving 100% improvement by Day 28. These clinical benefits support the potential for DBT-57A to improve the overall quality of life for diabetic patients. The clinical relevance of the product was further reinforced by a 95.24% improvement rate in most patients, although the 4.76% non-response rate indicates the need for further research to identify subgroups that may benefit more effectively. The high tolerability observed during the study, with only mild, self-limiting adverse effects reported, supports the continued use of DBT-57A under routine care. Supplementary laboratory analyses provided in S1 Table further confirm the product’s microbial safety and elemental composition within internationally accepted limits, reinforcing its suitability for observational evaluation.

While the findings demonstrate a significant reduction in fasting blood sugar levels following the use of DBT-57A, these results should be interpreted with caution. Given the observational nature of the study, the relationships observed are associative and do not imply direct causality. Future randomized controlled trials are needed to confirm causal efficacy.

#### 4.2 Proposed Mechanism of Action

While this study did not include pharmacological or biochemical testing to elucidate the exact mode of action of DBT-57A, the observed hypoglycemic effect is consistent with existing literature on the bioactivity of its constituent plants. Based on prior preclinical and pharmacological research findings, the following mechanisms are proposed:

The antidiabetic properties of DBT-57A are likely derived from the bioactive compounds found in its ingredients, such as quercetin, allyl disulphides, and glycoalkaloids. As a polyherbal product, the combination of these compounds with other secondary metabolites likely contributes to its observed blood sugar control. Quercetin and allyl propyl disulfide, compounds found in *Allium cepa* (onions), a key ingredient in DBT-57A, are known for their antidiabetic properties (28).

These compounds help lower blood sugar levels by enhancing insulin sensitivity and stimulating pancreatic *β*-cells to secrete insulin. Additionally, the antioxidant properties of these compounds reduce oxidative stress, improving overall metabolic health (29). Research has shown that *A. cepa* extracts reduce postprandial hyperglycaemia by inhibiting intestinal α-glucosidases, specifically sucrase and maltase, which delays digestion and glucose absorption (30). Furthermore, an ethyl alcohol extract of onion skin demonstrated antidiabetic effects comparable to acarbose, an α-glucosidase inhibitor, by delaying carbohydrate absorption and improving glucose homeostasis (31,32). Studies by Esakki et al. (2024) also reported that *Allium cepa* peels effectively inhibited α-glucosidase, decreased blood sugar levels, and enhanced insulin synthesis (33)

The leaves and bark extracts of *Khaya senegalensis*, have also demonstrated significant inhibitory activity against α-amylase and α-glucosidase in both in vitro and in vivo studies (34). This inhibition reduces blood sugar levels by slowing the breakdown of starches into glucose, In vivo studies have further shown that the bark extract effectively reduces blood sugar levels in a time-dependent manner (35).

*Solanum*, another ingredient of DBT-57A, has also been used traditionally to manage diabetes. Al-Ashaal and his colleagues found that the bioactive compounds-alkaloids and glycoalkaloids-in *Solanum torvum*, another ingredient in DBT-57A, has also been traditionally used to manage diabetes. The bioactive compounds alkaloids and glycoalkalooids present *Solanum torvum* exhibit significant antidiabetic activity. Studies have shown that these compounds inhibit hepatic glucose synthesis, enhancing peripheral glucose absorption and lowering blood sugar (36). In effect, the combination of therapeutic activities from *Allium cepa, Khaya senegalensis*, and *Solanum torvum*, gives DBT-57A its antidiabetic potential. The phytochemicals from DBT-57A likely exert their antidiabetic effects through a combination of insulin sensitization, glucose absorption inhibition, antioxidant activity, and anti-inflammatory actions. While these mechanisms support improved glycemic control, further research including biochemical testing and pharmacokinetic studies will be necessary to optimize dosages, refine formulations, and assess the long-term safety of DB-57A as a treatment for diabetes.

#### 4.3 Implications of the Study

This study underscores the critical importance of subjecting herbal medicines to structured scientific evaluation to ensure their safety, efficacy, and integration into evidence-based healthcare. The findings, which indicate a significant reduction in fasting blood sugar levels over a 28-day period suggest that DBT-57A hold potential as a supportive therapy in type 2 diabetes management. The observed rapid glycemic response also emphasizes the potential benefits of early intervention in mitigating long-term complications. Furthermore, these findings contribute to the growing body of evidence supporting the use of herbal medicines in modern healthcare. While these results are preliminary, they reinforce the need for regulatory engagement, public health advocacy, and interdisciplinary research to explore the therapeutic role of traditional herbal formulations within modern diabetes care models. Integrating such treatments into mainstream healthcare could provide alternative or complementary options for diabetes management, especially in resource-limited settings where access to conventional pharmaceuticals may be limited.

#### 4.4 Study Limitations and Future Research

While the findings of this study are promising, several limitations should be considered. This study is limited by its small sample size (25 participants), short duration (28 days), and lack of a control group, which affect the generalizability and causal interpretation of findings. Additionally, key health factors such as Body Mass Index (BMI), medication use, and diet were not analyzed, which may influence FBS changes. Future research should involve larger, randomized controlled trials with longer follow-ups, standardized symptom assessments, and mechanistic evaluations of insulin sensitivity and inflammatory markers to better understand the herbal mixture’s efficacy and safety. Also, additional factors influencing FBS improvement, including diet, physical activity, and medication adherence should be studied. Studies should also explore safety and efficacy parameters to ensure public safety and provide safer and effective alternative antidiabetic medicines.

### 5.0 Conclusion

This study provides preliminary evidence suggesting that DBT-57A may be an effective adjunct in the management of diabetes, demonstrating a significant reduction in fasting blood sugar levels and improvement in various diabetes-related symptoms. While the results are promising, they are based on a small sample size and short duration of observation. Larger, randomized controlled trials with extended follow-up are necessary to validate its clinical efficacy, assess long-term safety, and elucidate mechanisms of action. To ensure the safe and effective integration of DBT-57A into diabetes care, further studies should focus on the standardization of formulation, toxicological assessments, and pharmacovigilance. As interest in complementary and alternative medicine continues to grow, it is crucial to adopt a balanced approach that combines traditional knowledge with rigorous scientific evaluation to ensure the safe and effective management of diabetes.

## 5.2 Acknowledgement

The authors express their sincere gratitude to all the participants who volunteered for this study, as well as the clinical staff of the Health for All Herbal Clinic for their valuable support during data collection and follow-up. We also wish to acknowledge the contributions of the study’s research team; Dr. Edwin Laing, Prof. Oswald K. Seneadza, and Dr. Gabriel Okyere, whose efforts in the conceptualization and design of this study were instrumental to its successful execution.

## 5.3 Data Availability Statement

All relevant data supporting the findings of this study are included in the manuscript and its supplementary files. Due to ethical constraints related to patient confidentiality, de-identified individual-level data can be made available upon reasonable request from the corresponding author, subject to approval by the Committee on Human Research, Publications and Ethics (CHRPE), KNUST/Komfo Anokye Teaching Hospital.

